# Frequency of neutropenia over time in patients on clozapine

**DOI:** 10.1101/2022.01.25.22269860

**Authors:** Risha Govind, Amelia Jewell, Eromona Whiskey, Siobhan Gee, Ebenezer Oloyede, David Taylor, James H. MacCabe

**Affiliations:** Institute of Psychiatry, Psychology and Neuroscience, King’s College London, London, UK; South London and Maudsley NHS Foundation Trust, London, UK; National Psychosis Unit, South London and Maudsley NHS Foundation Trust, London, UK

**Author notes:** Corresponding author: James H. MacCabe.

## Abstract

**Background:** Clozapine, the only evidence-based drug for treatment-resistant schizophrenia is associated with agranulocytosis. For this reason, all clozapine patients are required to undergo mandatory regular blood monitoring throughout their clozapine treatment. The blood test results are reported using a traffic light system. The clozapine treatment is stopped immediately after a confirmed red result, which is the indication for risk of agranulocytosis. The need for blood tests places a burden on patients and acts as a barrier to clozapine treatment. There is growing evidence that the risk of agranulocytosis falls steeply after the first few months of treatment, raising the possibility that clozapine monitoring could be discontinued after a certain period of treatment.

**Aim:** To investigate the frequency density of the confirmed red results from clozapine monitoring across clozapine treatment.

**Method:** By merging electronic health records (EHR) data with clozapine blood monitoring data, we identified the clozapine treatment dates. The EHR data was from South London and Maudsley NHS Foundation Trust (SLAM). The clozapine blood monitoring data was from Zaponex Treatment Access System (ZTAS). ZTAS is one of the mandatory blood monitoring service providers in the United Kingdom. From these data, Kaplan-Meier survival curve was fitted to determine the time to get confirmed red results. At fixed points in the treatment, the future risk of obtaining a red result were calculated.

**Results:** By merging over 301,000 data points that came from the blood monitoring results and EHR data of 1,362 patients, we identified 1,891 clozapine treatment periods. Of these, 75 treatments were stopped due to confirmed red results. The Kaplan-Meier survival curve and the incidence rates data showed that 56 (74.7%) confirmed red results occur within the first 6 months of clozapine treatment.

**Conclusion:** We found a contrast between the relatively high density of the confirmed red results at the beginning of clozapine treatment which significantly reduces after 6 months of treatment which remained low thereafter.

## INTRODUCTION

Clozapine, the only evidence-based medication for treatment-resistant schizophrenia, is severely under-prescribed (1–3). This underutilization of clozapine is largely attributed to concerns regarding the risk of clozapine-induced agranulocytosis, an adverse drug reaction of clozapine that is prevalent in 0.4% of patients on clozapine (4–6).

Clozapine-induced agranulocytosis is a rare event with an unknown aetiology (7,8). Agranulocytosis, also known as severe neutropenia, is characterised as extremely low neutrophil count that results in increased susceptibility to fatal infections. Agranulocytosis is defined as a neutrophil count of <0.5×10^9^/L and neutropenia is defined as a neutrophil count of <1.5×10^9^/L. Clozapine, an atypical antipsychotic drug, was first introduced in Europe in 1971 for treating patients with schizophrenia (9). A few years later, clozapine was removed from the market after its use was shown to be associated with agranulocytosis (10). A seminal study by Kane et al in 1988 demonstrated its superior efficacy in treatment-resistant schizophrenia and led to the reintroduction of clozapine (1). However, in the United Kingdom (UK), United States and many other nations, clozapine use is subject to mandatory full blood count monitoring for the entire duration of clozapine treatment.

Under the current monitoring regulations in the UK, the blood monitoring starts with a baseline test and the frequency of the blood monitoring decreases with the length of clozapine treatment. A ‘baseline test’ refers to the monitoring test performed before clozapine treatment is started. The purpose of the baseline test is to ensure that the blood test results are stable before clozapine treatment can be allowed to be initiated. Once clozapine treatment is started, the patient is required to have their full blood counts monitored every week for 18 weeks, as shown in Box 1 (11). If the blood counts are stable in this period, then from 18 to 52 weeks, the blood monitoring is reduced to fortnightly. If the blood counts continue to be stable after 52 weeks of clozapine treatment, then blood monitoring is reduced to 4-weekly, but cannot be discontinued unless the patient stops taking clozapine (12). In the UK, the white blood counts (WBC) and absolute neutrophil count (ANC) are used to classify the results as either green, amber, or red (13). The patient’s further management is guided by this classification (Box 2).

Clozapine treatment must be stopped after a confirmed red result. This means that in the blood monitoring process, as soon as a red result is reported, a follow-up blood test is arranged. The clozapine treatment is stopped immediately if the follow-up blood test results is also red, thus confirming the initial red result. In the UK and Ireland, there is a ‘Central Non-Rechallenge Database’ to register patients who have had confirmed red results (14). The ANC for red results is < 1.5×10^9^/L, which is the definition for neutropenia, therefore red results can be used to indicate neutropenia.

The mandatory blood monitoring throughout the clozapine treatment is a major burden to patients and health services, and contributes to the underutilisation of clozapine (15). In this study, we studied the frequency density of the confirmed red results over time using the results from the mandatory blood monitoring.

### Box 1

**UK clozapine monitoring frequency**

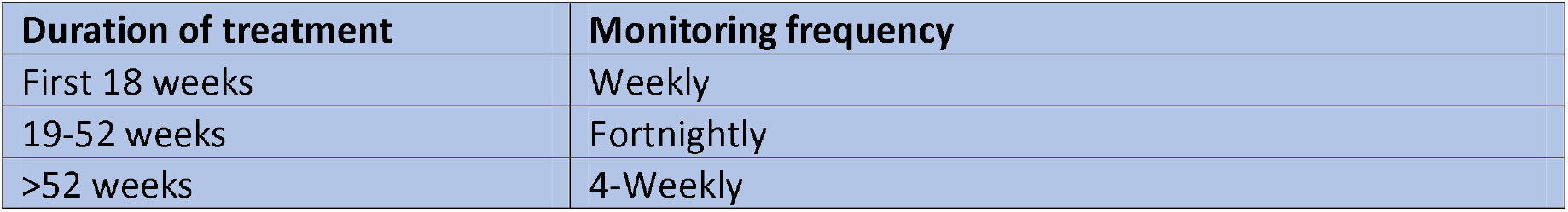

**Box 1: UK clozapine monitoring frequency**
| Duration of treatment | Monitoring frequency |
| --- | --- |
| First 18 weeks | Weekly |
| 19-52 weeks | Fortnightly |
| >52 weeks | 4-Weekly |

### Box 2

**UK clozapine monitoring results classification criteria**

**Box 2: UK clozapine monitoring results classification criteria**
| Classification | WBC<br>($\times 10^9/L$) | ANC<br>($\times 10^9/L$) | Guidance |
| --- | --- | --- | --- |
| ● Green | >3.5 | >2.0 | Continue treatment |
| ● Amber | 3.0-3.5 | 1.5-2.0 | Continue, but monitor twice weekly until green results |
| ● Red* | <3.0 | <1.5* | Arrange emergency blood test to confirm red result.<br>If red result is confirmed, then STOP TREATMENT |

\*: ANC < $1.5 \times 10^9/L$ is also the definition of neutropenia. Therefore, red results can be used indicate neutropenia.

## METHOD

### Data sources

#### CRIS

The Clinical Record Interactive Search (CRIS) is a database containing the fully de-identified health records of SLAM. SLAM caters to all secondary mental health care needs of over 1.3 million people of four London boroughs (Lambeth, Southwark, Lewisham, and Croydon); CRIS provides the platform for all their electronic health records to become available to researchers for secondary analysis within a robust data security and governance framework (16).

CRIS is comprised of both structured and free-text fields from the SLAM’s clinical notes. In this study, two types of data were extracted from CRIS, pharmacy dispensary data and clozapine clinic attendance data.

The pharmacy dispensary data included the strength and quantity of clozapine that was dispensed to the patient. This data was available in the structured fields. The earliest record of pharmacy data used in this study was from September 2005.

The clozapine clinic attendance data came from the nurse’s record of the patient attending the appointment for their routine clozapine blood test. This information was retrieved by combining information from free-text fields as well as structured fields. The free-text fields were searched for the phrases “clozapine clinic” or “clozaril clinic”. The structured fields were searched for the entry “attended”. The dates on which both the components were retrieved became the clozapine clinic attendance data. The two phrases, “clozapine clinic” or “clozaril clinic” were selected based on exploring the free-text notes and finding that even though the brand Zaponex is used at SLAM, some healthcare providers who write clinical notes tend to refer to refer to clozapine using the brand name ‘clozaril’. The earliest record of clozapine clinic attendance data used in this study was from November 2002.

#### ZTAS

Zaponex Treatment Access System (ZTAS) is one of the UK’s mandatory blood monitoring service providers. Clozapine patients treated at South London and Maudsley NHS Foundation Trust (SLAM) have their blood counts monitored by ZTAS (http://www.ztas.co.uk).

The ZTAS data was available via the SQL Server Management Studio version 15.0 (Microsoft Inc, USA). In three separate SQL database tables, ZTAS stores detailed information on the clozapine blood monitoring results (described further below), detailed information on clozapine treatment statuses (described further below) that were recorded, and clozapine treatment start dates (described further below.

The blood monitoring SQL database table of ZTAS includes the date, white blood counts (WBC), absolute neutrophil counts (ANC) and the result classification of each blood test records. It also includes labels for blood tests that were a baseline. Box 2 gives details of how the results are classified. The classifications of the results are either green, amber and red. Green results means continue treatment, amber means increase monitoring and red means to reconfirm this result, and a confirmed red result requires the immediate discontinuation of clozapine treatment.

The treatment status SQL database table of ZTAS includes the date and status label for all recorded updates in the status of clozapine treatments. The status labels can change multiple times for the same patient. Examples of the statuses are ‘on-treatment’, ‘interrupted’, ‘discontinued’, ‘transferred’ and ‘non-rechallengable’.

The start status SQL database table of ZTAS contained only one clozapine treatment start date per patient. This date was the closest treatment start date prior to the date the linkage was made to access ZTAS data via the CRIS platform (described further below).

All data from these three SQL database tables that were accessible via CRIS were used in this study.

#### Linkage and cohort definition

A linkage was made from the CRIS database to the ZTAS database. The linkage was performed by mapping patient identifiers from CRIS (including name, NHS number, date for birth) to those in the ZTAS data, and then pseudo-anonymising the data, thus making it seamless to perform secondary analysis using the ZTAS data while patient identities remain de-identified (17,18).

The databases were initially linked in March 2016 and the linkage was updated in October 2019, therefore even though ZTAS continues to monitor SLAM patients, our dataset includes only those who were on both databases on either or both of the linkage dates. This comprised of almost 20 years of ZTAS data, ranging from 2^nd^ May 2000 to 1^st^ October 2019, inclusive.

All CRIS data were extracted until the date of the last record available from ZTAS, 1^st^ October 2019. The cohort comprised all SLAM patients who were in the ZTAS linkage database in CRIS.

### Identification of clozapine treatment start dates

SQL Server Management Studio version 15.0 (Microsoft Inc, USA) was used to extract the data and standard Python (V.3.7.4) libraries were used for identifying the clozapine treatment dates.

The clozapine treatment dates that were provided by ZTAS only included one start date per person, and this was the closest start date prior to the date of linkage with CRIS data. In order to find all the treatment start dates, we developed an algorithm using data from ZTAS and CRIS. The data from ZTAS included clozapine blood monitoring results, clozapine treatment statuses and clozapine treatment start dates. The data from CRIS included the pharmacy dispensary data and the clozapine clinic attendance data. Figure 1 shows the study flow chart.

**Figure 1:**
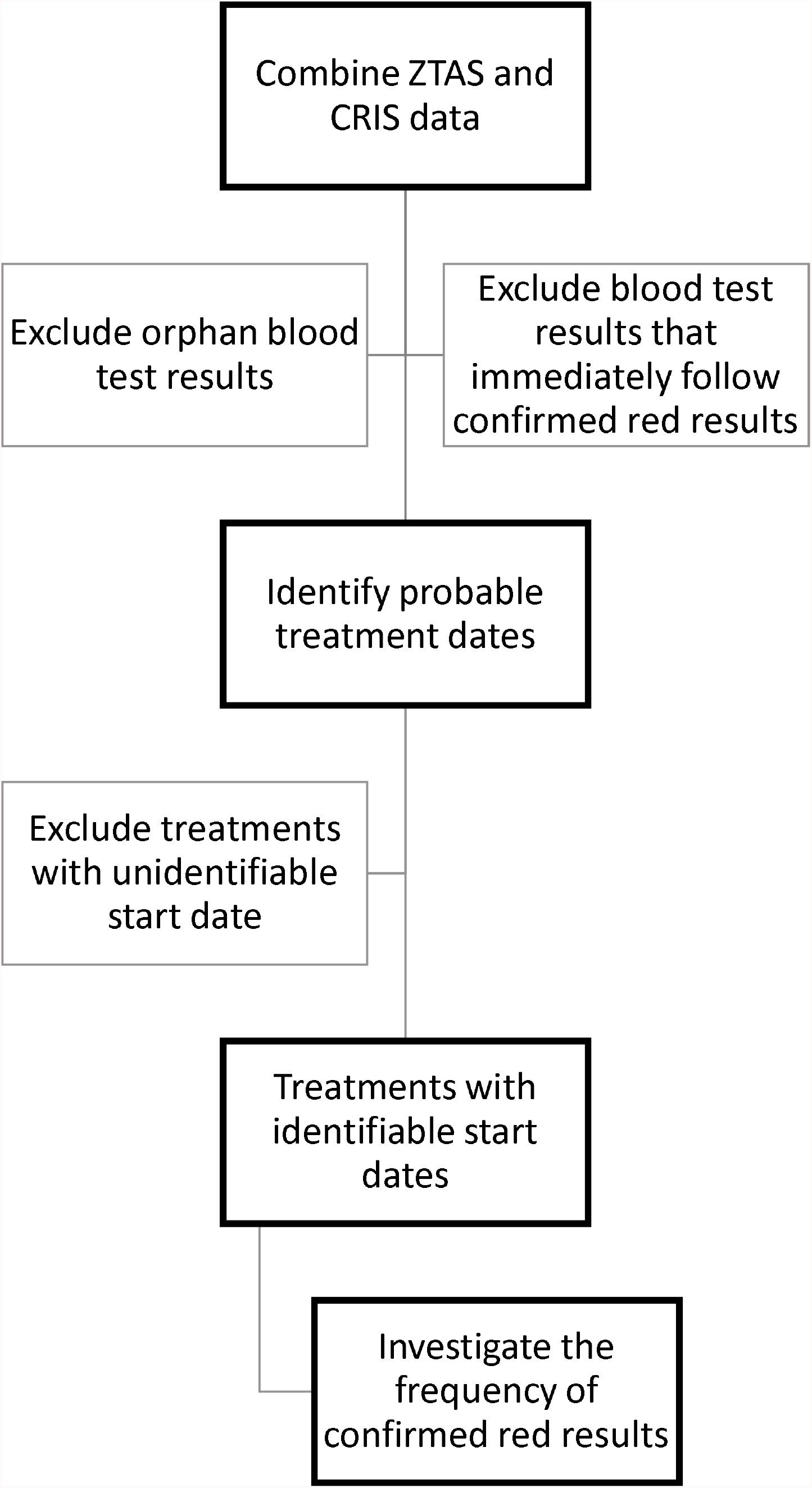
Study Flow chart

#### Data Cleaning

The first step of data cleaning was to remove orphan blood tests from the analysis. A baseline test is mandatory before the clozapine treatment is started. Therefore, if a single test exists with no subsequent blood tests, then it can be assumed that that the clozapine treatment was not started at that time. We defined an orphan blood test as a single blood test with no blood tests within 60 days, prior or subsequent to it. The 60 days threshold was used to accommodate for 1 missing datapoint since most of the tests are at 4-weekly frequency.

The second step of data cleaning was to remove the blood test data that occurred immediately after the clozapine treatment was stopped due to a confirmed red result, so that only blood tests on clozapine treatment were included. This criteria was created on the basis that clozapine treatments are stopped as soon a confirmed red result occurs. It is common for the blood monitoring to continue after the clozapine treatment is stopped due to confirmed red results until the patient’s blood results are back in the green zone (Box 2).

#### Probable treatment dates

Using the cleaned data, we identified the probable start and end dates of clozapine treatment.

A probable start date of clozapine treatment was identified if any of these 3 criteria were fulfilled:

1. The blood test was the first blood test result of a patient in our dataset
2. The presence of a ‘baseline’ label on blood test result
3. The blood test was after a gap of >35 days for short treatments (see below for definition) or a gap of >60 days for long treatments (see below).

A treatment was classified as ‘short’ if it was up to 18 weeks long, and ‘long’ if it was longer than 18 weeks.

A probable end date of clozapine treatment was identified if any of these 4 criteria were fulfilled:

1. The blood test was the last blood test result of a patient in our dataset
2. The presence of a confirmed red result
3. The blood test occurred immediately prior to a gap of >35 days for short treatments or >60 days for long treatments
4. The ZTAS status changed to ‘discontinued’, ‘interrupted’ or ‘transferred’.

Once the probable treatment dates were identified, we performed an additional quality control check by examining the frequency of blood tests around each probable start date and end date. Any probable start dates, and probable end dates appearing in the middle of an on-going treatment period and followed by tests at a frequency other than weekly, were removed and not used in the analysis.

#### Treatments with identifiable start dates

Where clozapine treatment was started when the patient was under the care of a different healthcare provider, and the patient was later transferred to SLAM, we had missing data regarding the start date of the clozapine treatment. For these treatments, the probable start dates that we identified were pointing to the time the clozapine treatment started in SLAM instead of the actual start of the clozapine treatment. Therefore, treatments without identifiable start dates were removed. Since it is mandatory for all clozapine treatments to start with a baseline test, followed by weekly blood tests for the first 18 weeks, a treatment had an identifiable start date if the first test of the treatment had a baseline label AND the mode or mean of the frequency of tests in the first 18 weeks was ≤ 7 days.

To cater for the possibility of missing baseline labels, two additional requirements needed to be fulfilled if the first test of the treatment did not have a baseline label. The additional requirements for these treatments were that the median or mean of the frequency of the tests between 13^th^ to 18^th^ week of treatment was ≤ 7 days AND if the treatment was longer than 18 weeks, then there were >15 tests in the first 18 weeks of treatment.

To cater for treatments that would come under of category of ‘off-licence’, treatments that started any time after the patient had a confirmed red result and therefore would have been in the non-rechallengable register needed to fulfil an additional requirement. ‘Off-licence’ refers to the use of clozapine outside the marketing authorisation issued in the UK for the drug, meaning the patient is receiving clozapine treatment after being registered register as non-rechallengable (19). The additional requirement for these treatments was for their ZTAS status to change from ‘non-rechallegable’ to ‘on-treatment’. Note that these treatments were not comprised of the tests that immediately followed a confirmed red result as those blood tests was already removed earlier in the analysis.

The treatments with unidentifiable start date that ended due to confirmed red results were manually checked to confirm that it was indeed not possible to identify the start dates of these treatments within the scope of the existing data.

### Statistical Analysis

The statistical analysis was performed using STATA for Windows version 15.1. A Kaplan-Meier survival curve was fitted to display the time to confirmed red results. The confirmed red results were used as the failure event in this analysis.

The stptime command of STATA was used to compute and tabulate the person-years and incidence rates of confirmed red results against the length of clozapine treatment. The person-years is the sum of the number of years each patient has been on treatment. The incidence rates refer to the number of confirmed red results divided by the person-years. Due to the low incidence of confirmed red results in certain period of treatment, the incidence rates were measured per 1,000 person-years.

Using the person-years and number of confirmed red results data, the future incidence rates of red results at different points of clozapine treatment were calculated. First, the future person-years were calculated by finding the sum of the number of years of treatments remaining at specific point in treatments. These future incidence rates were calculated by dividing the future number of confirmed red results by the future person-years at specific points in clozapine treatment. The future incidence rates were also measured per 1,000 person-years. This calculation was performed using Microsoft Excel version 2102 (Microsoft Inc, USA).

### Ethical considerations

CRIS was approved for use as a de-identified data resource for secondary analysis by Oxfordshire Research Ethics Committee C (reference 18/SC/0372).

## RESULTS

Between 2^nd^ May 2000 and 1^st^ October 2019, ZTAS recorded 210,273 blood test results for 2,028 SLAM patients. The number of blood tests per person ranged from 1 to 341. The median number of tests per person was 94.

Figure 2 shows that after removing orphan blood tests and blood tests that were performed immediately after the confirmed red result, there were 208,554 tests remaining. These came from 1,988 SLAM patients and comprised of 3,167 probable treatment periods.

**Figure 2:**
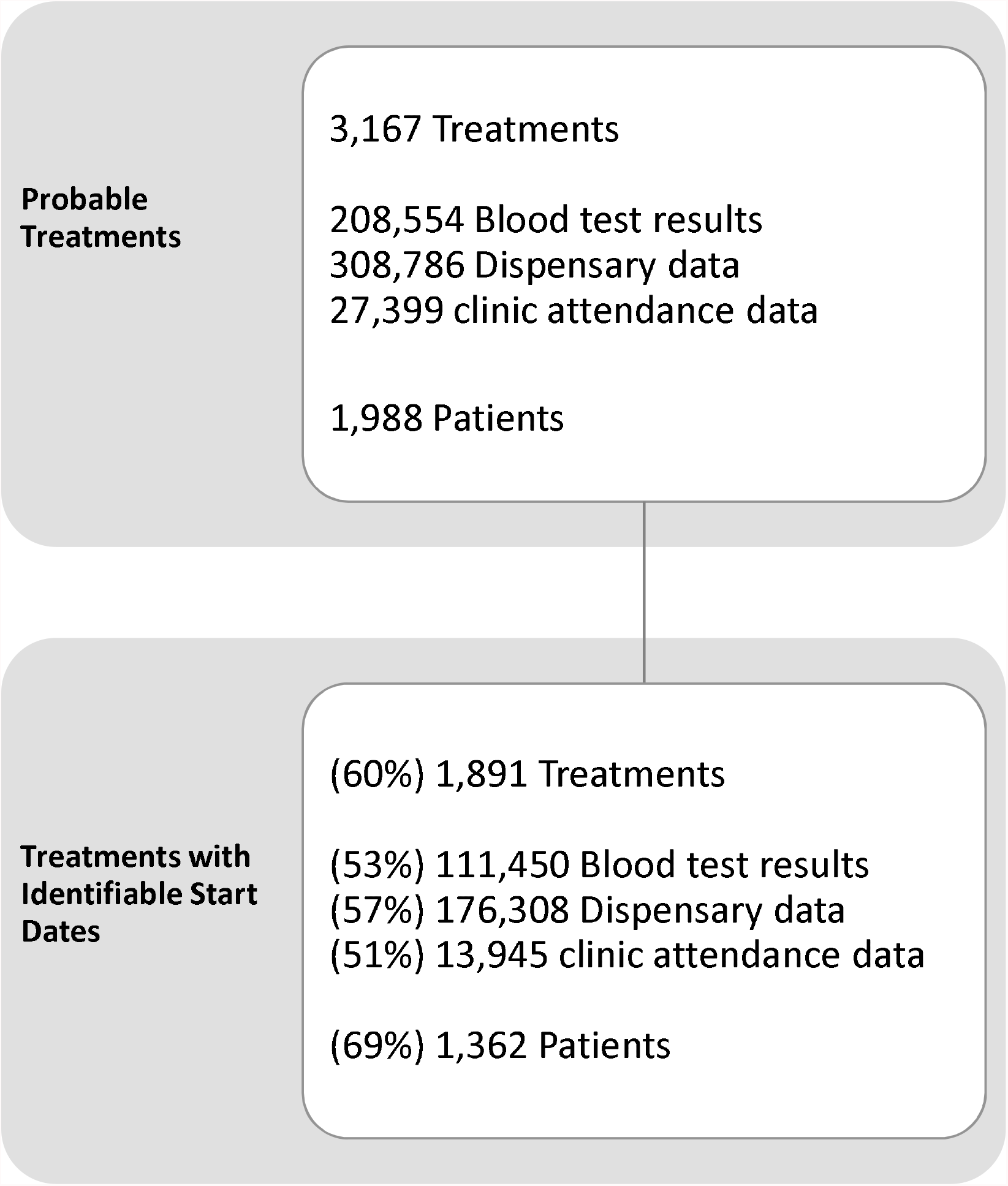
Figure showing the numbers of probable treatment periods identified, of these, the treatment periods with identifiable start dates. Treatment periods were calculated from combining blood test results, dispensary data and clinic attendance data. Not all treatments had identifiable start dates due to missing data, as those treatments started in other settings.

Of the 3,167 probable treatments, 1,276 (40%) had an unidentified start date, and were thus excluded from the analysis. 24 (1.9%) of the treatments with unidentifiable start dates ended in confirmed red results. These 24 treatments were manually checked, and this confirmed that all these treatments started outside the scope of the data available.

Of the 3,167 probable treatments, we were able to identify the start dates of 1,891 (60%) of them, of which 1,551 (82%) had their first test labelled as ‘baseline’. The treatment lengths ranged from 2 days to 19 years, with a median of 1.1 years. 75 of these treatments ended with the confirmed red results. The majority (74.7%) of the treatments that ended due to confirmed red results ended within the first 6 months of treatment.

Figure 3 shows the distribution of the confirmed red results over time. This plot was generated using the 75 treatments that ended with confirmed results with known start dates. It shows that majority of the confirmed red results occur in the first 6 months of treatment. After 6 months, the incidence of confirmed red results is sporadic. This pattern is reflected in the Kaplan Meier survival analysis and in the incidence rates the future incidence.

**Figure 3:**
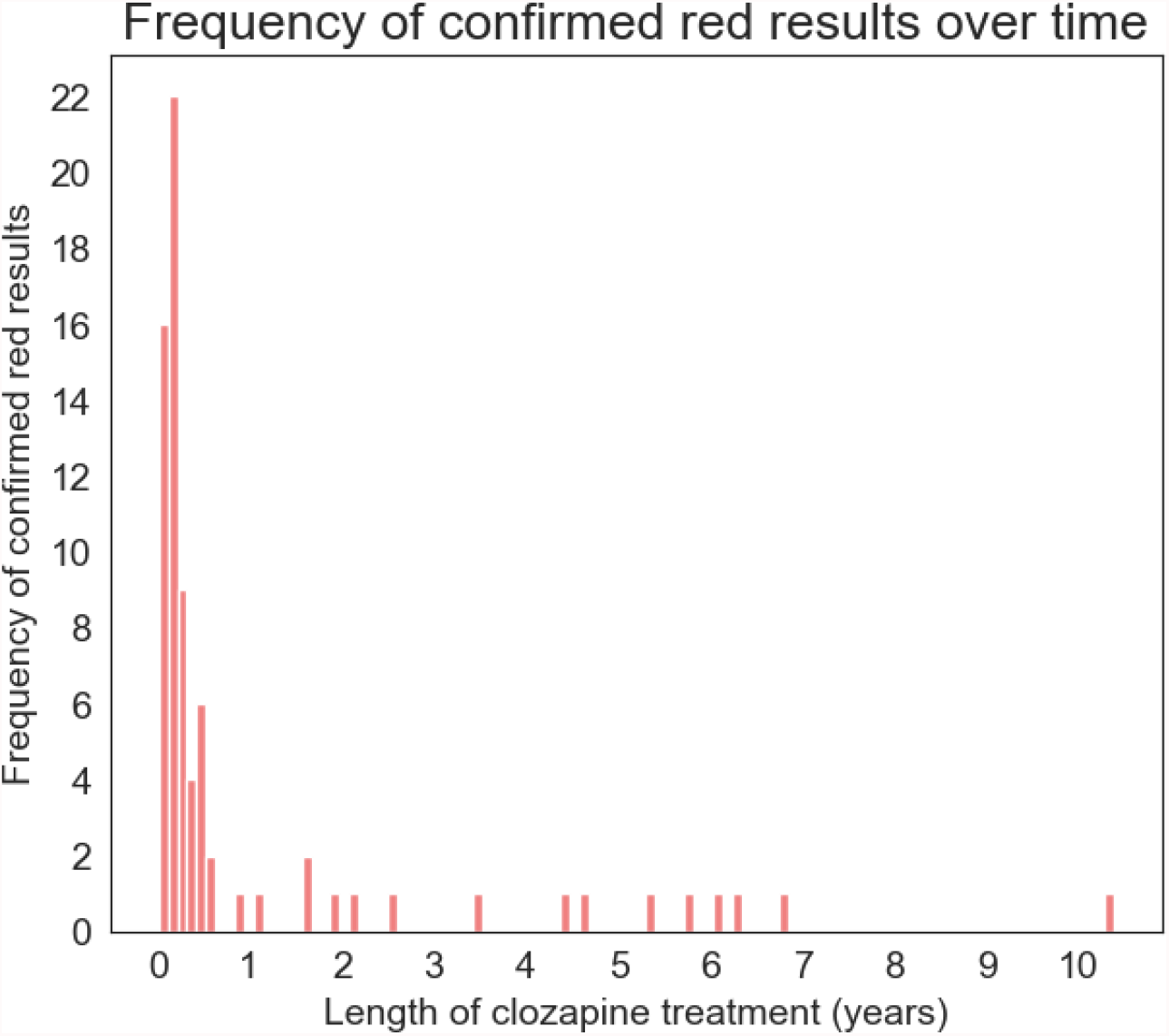
The Histogram of confirmed red results over time plot shows that majority of the confirmed red results occur in the first 6 months. After 6 months of treatment, the incidence of confirmed red results is sporadic. This pattern is reflected in the Kaplan Meier survival analysis and in the incidence rates the future incidence. One explanation of this pattern could be that there are two distinct biological mechanisms, and the clozapine-induced immunological response occurs in the first 6 months of clozapine treatment and afterwards, the occurrence of the confirmed red results is random.

Figure 4 shows the Kaplan-Meier survival plot of the length of clozapine treatment to get confirmed red results. This plot was generated using the 1,891 treatments that had known start dates. Of the 75 treatments that ended with confirmed red results, 56 (74.7%) confirmed red results occur in the first 6 months of clozapine treatment. The plot demonstrates 3 distinct phases of risk for getting a confirmed red result: (I) the risk is highest in the first 6 months of treatment (II) the risk reduces to a reasonably constant level from after 6 months to 7 years of treatment (III) after 7 years the risk is almost zero as, after 7 years, there is only 1 incidence of a confirmed red result, and that is at 10.4 years.

**Figure 4:**
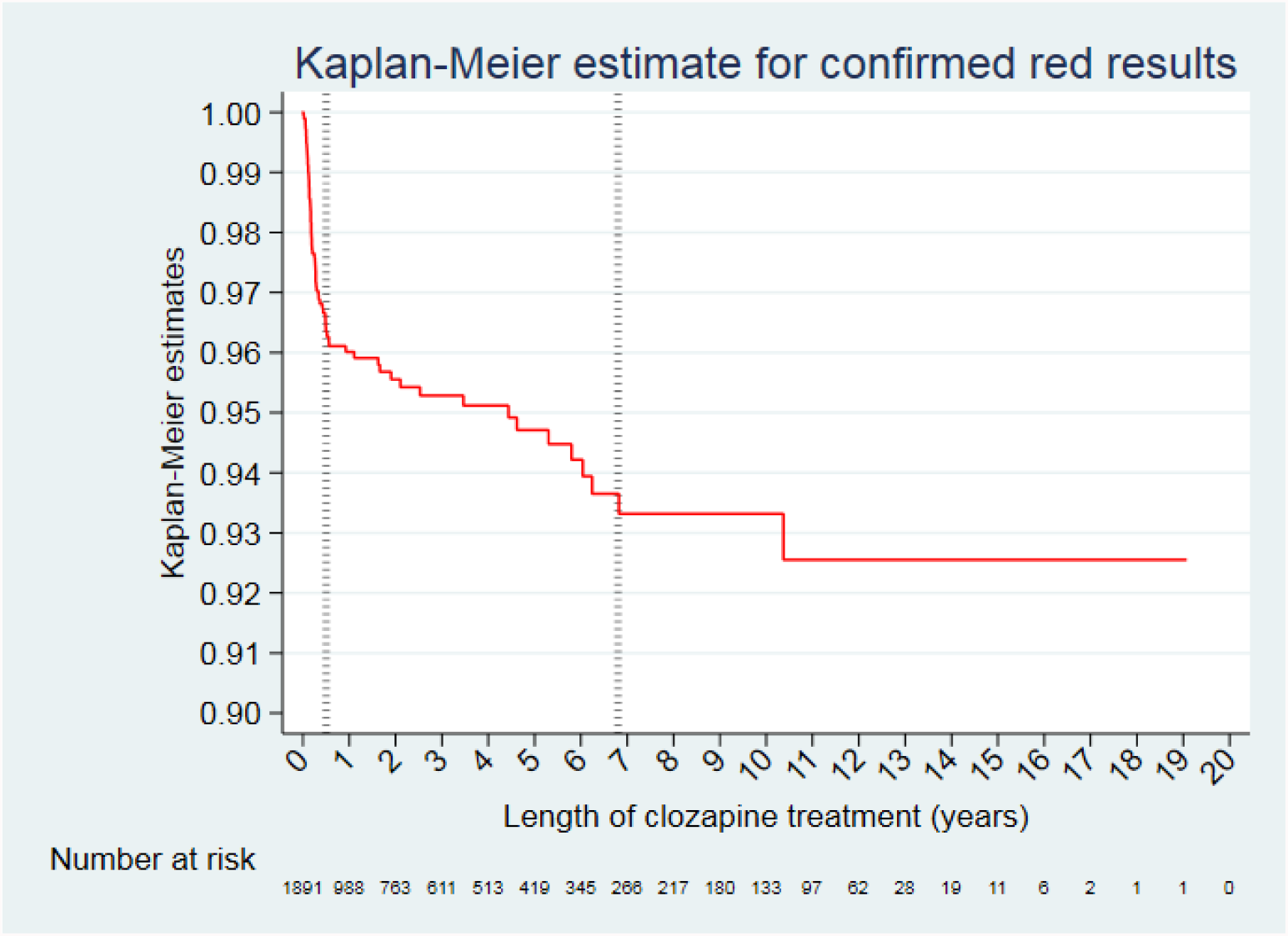
Kaplan-Meier plot of confirmed red results. Below the plot shows the number at risk at each year. The plot demonstrates 3 distinct phases of risk for getting a confirmed red result: (I) the risk is high in the first 6 months of treatment (II) the risk is low and reasonably constant from after 6 months to 7 years of treatment (III) after 7 years the risk is almost zero as, after 7 years, there is only 1 incidence of a confirmed red result

Table 1 shows the person-years and incidence rates of confirmed red results at different time points in clozapine treatment. Compared with the rest of the treatment, the first year of treatment has a higher incidence rate of confirmed red results, 47 (95% CI: 36-60) per 1,000 person-years. The incidence rate of confirmed red results in the second year of treatment is 5 (95% CI: 2-12) per 1,000 person-years. The majority of confirmed red results occur in the first 4 months of clozapine treatment. The overall incidence rate of confirmed red results at any time in the treatment is 14 (95% CI: 11-17) per 1,000 person-years.

**Table 1:** Number of confirmed red results, person-years, and incidence rates of the confirmed red results as clozapine treatment progresses, shown for every month for the first 12 months, then for every year. Person-years is the sum of years in all the treatments that ended within the specified period. The incidence rate for each period is refers to the number of confirmed red results divided by the person-years. The incidence rates are displayed per 1,000 person-years.

| Length of clozapine treatment | Number of confirmed red results in this period | person-years | Incidence Rate per 1,000 person-years (95% CI) |
| --- | --- | --- | --- |
| 0-1 years | 60 | 1284.5 | 47 (36-60) |
| 0-1 months | 10 | 147.1 | 68 (37-126) |
| 1-2 months | 17 | 132.6 | 128 (80-206) |
| 2-3 months | 11 | 122.6 | 90 (50-162) |
| 3-4 months | 9 | 116.3 | 77 (40-149) |
| 4-5 months | 3 | 110.2 | 27 (9-84) |
| 5-6 months | 6 | 105.3 | 57 (26-127) |
| 6-7 months | 3 | 100.1 | 30 (10-93) |
| 7-8 months | 0 | 96.9 | 0 |
| 8-9 months | 0 | 93.4 | 0 |
| 9-10 months | 0 | 90.0 | 0 |
| 10-11 months | 0 | 86.4 | 0 |
| 11-12 months | 1 | 83.6 | 12 (2-85) |
| 1-2 years | 4 | 857.1 | 5 (2-12) |
| 2-3 years | 2 | 683.9 | 3 (1-12) |
| 3-4 years | 1 | 558.0 | 2 (0-13) |
| 4-5 years | 2 | 468.2 | 4 (1-17) |
| 5-6 years | 2 | 385.7 | 5 (1-21) |
| 6-7 years | 3 | 304.4 | 10 (3-31) |
| 7-8 years | 0 | 240.4 | 0 |
| 8-9 years | 0 | 197.1 | 0 |
| 9-10 years | 0 | 156.6 | 0 |
| 10-11 years | 1 | 115.9 | 9 (1-61) |
| 11-12 years | 0 | 78.5 | 0 |
| 12-13 years | 0 | 43.8 | 0 |
| 13-14 years | 0 | 22.1 | 0 |
| 14-15 years | 0 | 14.4 | 0 |
| 15-16 years | 0 | 9.1 | 0 |
| 16-17 years | 0 | 3.9 | 0 |
| 17-18 years | 0 | 1.9 | 0 |
| 18-19 years | 0 | 1.0 | 0 |
| 19-20 years | 0 | 0.1 | 0 |
| total | 75 | 5426.6 | 14 (11-17) |

Table 2 shows the future person-years and incidence rates of confirmed red results at different time points in clozapine treatment. The rate of confirmed red results are the highest at the beginning of treatment. The future incidence rate of confirmed red results in the 1^st^ month of treatment is 13.8 per 1,000 person-years. This rate gradually decreases until the 6^th^ month of treatment, where the future incidence rate of confirmed red results is 4.0 per 1,000 person-years. The future incidence rate remains below 4.0 for the rest of the treatment.

**Table 2:** Table of the incidence rate (per 1,000 person-years) for getting a red result in at different time points in clozapine treatment. This statistic was calculated by dividing the future number of confirmed red results by the future person-years at specific points in clozapine treatment. The future person-years is the sum of the number of years of treatments remaining at specific point in treatments.

| Point in clozapine treatment | Future number of confirmed reds | Future person-years | Future Incidence Rate per 1,000 person years |
| --- | --- | --- | --- |
| 0 months | 75 | 5426.6 | 13.8 |
| 1 month | 65 | 5279.5 | 12.3 |
| 2 months | 48 | 5146.9 | 9.3 |
| 3 months | 37 | 5024.3 | 7.4 |
| 4 months | 28 | 4908.0 | 5.7 |
| 5 months | 25 | 4797.8 | 5.2 |
| 6 months | 19 | 4692.5 | 4.0 |
| 7 months | 16 | 4592.4 | 3.5 |
| 8 months | 16 | 4495.5 | 3.6 |
| 9 months | 16 | 4402.1 | 3.6 |
| 10 months | 16 | 4312.1 | 3.7 |
| 11 months | 16 | 4225.7 | 3.8 |
| 1 year | 15 | 4142.1 | 3.6 |
| 2 years | 11 | 3285.0 | 3.3 |
| 3 years | 9 | 2601.1 | 3.5 |
| 4 years | 8 | 2043.1 | 3.9 |
| 5 years | 6 | 1574.9 | 3.8 |
| 6 years | 4 | 1189.2 | 3.4 |
| 7 years | 1 | 884.8 | 1.1 |
| 8 years | 1 | 644.4 | 1.6 |
| 9 years | 1 | 447.3 | 2.2 |
| 10 years | 1 | 290.7 | 3.4 |
| 11 years | 0 | 174.8 | 0 |
| 12 years | 0 | 96.3 | 0 |
| 13 years | 0 | 52.5 | 0 |
| 14 years | 0 | 30.4 | 0 |
| 15 years | 0 | 16.0 | 0 |
| 16 years | 0 | 6.9 | 0 |
| 17 years | 0 | 3.0 | 0 |
| 18 years | 0 | 1.1 | 0 |
| 19 years | 0 | 0.1 | 0 |
| 20 years | 0 | 0 | 0 |

## DISCUSSION

### Summary of findings

We investigated if risk of clozapine-induced neutropenia changes with clozapine treatment duration using the clozapine blood monitoring data. We used the confirmed red results as the indication of neutropenia. We found a contrast between the relatively higher risk at the beginning of clozapine treatment and significantly reduced after 6 months of treatment.

### Comparison with previous studies

To our knowledge, no previous research has specifically investigated the risk of neutropenia using clozapine blood monitoring data. Our results are consistent with the findings of Amsler et al in 1977 where they reported that all their observed cases of severe neutropenia occurred during the first 3 months of clozapine (20).

### Strengths and limitations

As a strength of this study, this study was based on SLAM patients. SLAM has a near-monopoly in providing all aspects of secondary mental health care to over 1.3 million people of four London boroughs (Lambeth, Southwark, Lewisham, and Croydon), thus creating an ascertainment to study the effects of antipsychotics drugs, such as clozapine on patients in the UK.

The main limitation of this study is missing data. The study was performed on data available to us from the electronic health records of SLAM and the clozapine blood monitoring data from ZTAS, latter was used as the primary data for identifying the dates of clozapine treatment. Unfortunately, we could not identify the start dates of 40% of the treatment periods, which comprised of 47% of blood monitoring data. These clozapine treatments started prior to the date of first blood monitoring data we had of these patients. ZTAS became SLAM’s blood monitoring service provider in the year 2000. Since we only have access to the blood monitoring results of one clozapine monitoring service, ZTAS, we were not able to identify their start dates of clozapine treatments that started pre-2000. Similarly, we were not able to identify the start dates of patients who transferred from another trust where they started their clozapine treatment. The treatments with unidentifiable start dates were removed from the analysis, and this included 24 treatments that ended with confirmed red results. We manually checked the data on these 24 treatments to confirm that they start dates were unidentifiable using the data available to us.

Another limitation was that although the information on clozapine start dates were embedded in the free-text clinical notes, extracting this information was a major challenge. Because of the lack of standards in writing the clinical notes, embedded information within the clinical notes is currently not utilized to its fullest potential in research in general(21).

The findings on future incidence of red results should be treated with caution. Some of these results, particularly for later in treatment, are based on low numbers. However, we believe these data should be presented as they make the general point that the future risk of red results (on which mandatory testing is predicated) is substantially lower in patients with long periods of treatment.

### Implications

To our knowledge, our study is the first to present data on the risk of having a future confirmed red result and that the risk falls to low levels after 1 year. The risks of a future red result should be weighed against the risks and burdens of monitoring, including the risk of psychotic relapse due to unnecessarily discontinuing clozapine due to a low neutrophil count that is unrelated to clozapine treatment. This supports the case for reducing the monitoring even further as treatment progresses because as treatment progresses, the risk-benefit ratio of monitoring changes significantly. It also suggests that the rigid application discontinuation rules based on thresholds may not be appropriate, at least after 6 months. An alternative system could be proposed, whereby, if a confirmed red result is obtained after 6 months, a haematological review could be triggered to try to determine whether the cause of the neutropenia was likely to be related to clozapine or not, and to advise the treating team on the likely risks of clozapine rechallenge.

### Future Work

Since with the current methods, we were only able identify the start dates of just 53% of the treatments, we will be exploring computational solutions such as Natural Language Processing (NLP) to identify more clozapine start dates from information embedded in the free-text clinical notes. Also, we will perform cost-effectiveness analysis to model the tipping point where the benefit of monitoring no longer outweighs the burden and cost of clozapine monitoring to the healthcare system as well as the patient.

## Data Availability

This is a retrospective cohort study was carried out using data from the electronic records of the South London and Maudsley NHS Foundation Trust (SLAM). This data cannot be made available. The analysis used a de-identified data resource approved by Oxfordshire Research Ethics Committee C (reference 18/SC/0372).

## Ethics statement

The research was conducted under ethical approval reference 18/SC/0372 from Oxfordshire Research Ethics Committee C.

